# Development and validation of the Dysarthria Impact Scale

**DOI:** 10.1101/2025.07.02.25330604

**Authors:** Adam P. Vogel, Lisa Graf, Merit Gauß, Cheuk S. J. Chan, Graham Hepworth, Matthis Synofzik

## Abstract

**Background:** Impaired speech due to dysarthria significantly impacts quality of life. Patient-reported outcomes (PROs) offer critical insight into the lived experience of communication disability and are increasingly central to regulatory frameworks for patient-focused drug development.

**Objectives:** To develop and validate the Dysarthria Impact Scale (DIS), a brief PRO designed to assess the impact of motor speech disorders on quality of life across neurological conditions.

**Methods:** A multi-site, cross-sectional study was conducted with 244 participants, including individuals with Huntington’s disease, Parkinson’s disease, hereditary ataxias, and head and neck cancer, along with healthy controls. The 22-item DIS was developed using expert input and patient feedback and evaluated alongside reference tools (Voice Handicap Index and SF-36). Item reduction procedures yielded two shorter versions (DIS-17 and DIS-6). Validity, reliability, and sensitivity/specificity analyses were performed, and minimal clinically important differences (MCIDs) were estimated using distribution-based methods.

**Results:** All DIS versions showed strong convergent validity with the VHI (r = –0.85) and SF-36 (r = 0.72) and were correlated with blinded perceptual speech ratings. DIS-17 and DIS-6 achieved comparable sensitivity (0.93 and 0.88) and specificity (0.84 and 0.86, respectively). Test–retest reliability was high (r = 0.98), with WSSD estimates ranging from 4.0 to 10.6 across groups. Estimated MCIDs ranged from 5 to 15 points. Group differences were observed, with lower DIS scores in ataxia and Parkinson’s disease compared to Huntington’s disease.

**Conclusions:** The DIS is a valid, reliable, and practical PRO for quantifying the impact of dysarthria on quality of life. It is suitable for clinical monitoring and clinical trial use, with sensitivity to disease-related variation and change.

## Introduction

Brain disease or injury causing motor dysfunction frequently results in disordered speech. This impairment can lead to daily disadvantage and stigmatization ^1-3^. Dysarthria is the most common neurological speech disorder. It is characterized by sensorimotor impairment in one or multiple speech subsystems including respiration, phonation, articulation, resonance, and prosody. Deficits in any or all these subsystems can reduce intelligibility (ability to be understood) and naturalness (deviation from a healthy norm) and impact overall communicability. It is established that communication deficits trigger altered self-identity ^4^, impede social ^5^ and professional interactions ^3^, and lead to social marginalization ^6^. Seventy percent of people with a communication disorder are unemployed or in the lowest income brackets ^7^. These conditions have profound health, psychosocial and economic consequences yet tools to evaluate the impact of dysarthria on patients are limited.

The nature and impact of dysarthria on the speaker varies based on factors including site, size and type of lesion or neurological damage. For example, neurodegenerative diseases like Huntington’s disease, Parkinson’s disease or hereditary ataxia lead to a progressive decline of imprecise production of consonants, vowel distortion, altered resonance and slower rate of speech ^8-10^. Whereas stroke or traumatic brain injury or neurodevelopmental conditions (e.g., FOXP2) may present with static deficits across subsystems ^11-14^. Both neurodegenerative and non-progressive etiologies appear to be amenable to treatment ^15-22^; however there are no pharmaceutical therapies specifically designed to improve speech.

Tools specifically built for measuring dysarthria related quality of life available to clinicians and researchers include the Dysarthria Impact Profile ^23^, the QOL-DyS ^24^, and the Living with dysarthria: evaluation of a self-report questionnaire ^25^. These tools are limited by their scope, validation protocols, cohort make up, complexity of question design, length of survey and narrow language options beyond English. For example, some tools are validated across heterogenous clinical cohorts (e.g., Living with dysarthria), some have more than 100 items (e.g., Dysarthria Impact Profile, which is too long for many protocols, some were not validated against standardized comparators (e.g., ^24^) and some employ both negative and positive questioning (e.g., Dysarthria Impact Profile) which is confusing for individuals with and without a cognitive impairment.

A comprehensive assessment of motor speech impairment requires the use of patient reported outcomes (PROs) specifically focused on dysarthria related quality of life. Also, patient-focused drug development requires trial outcomes (e.g. of speech) to be anchored to outcomes shown meaningful to and by patients and their network. Put another way, the US Food and Drug Administration (FDA) describes clinical outcome assessments (COAs) as measures that directly quantify what matters most to individuals, including how they function and feel ^26, 27^. Consequently, assessment of any impact of treatment, either pharmaceutical or behavioral, or disease (either neurodegenerative or non-progressive) should include a combination of objective and subjective tests ^28^. In clinical trials, tests should be brief, repeatable and suitably motivating to ensure completion at regular intervals ^29, 30^. Here we describe development and validation of the Dysarthria Impact Scale, as a novel brief PRO for assessing the impact of dysarthria for people with motor speech impairment, to be used in clinical assessments, natural history studies or treatment trials.

## METHODS

### Tool development

A protocol for building the patient reported outcome was established prior to commencing the study. The COSMIN risk of bias checklist was used as a guide to optimize methodological quality and limit risk of bias of the proposed tool ^31, 32^. The *a priori* clinometric properties targeted in the tool design focused on integrating adequate reliability and validity, minimizing measurement error and maximizing responsiveness to symptom severity. It was considered important that the proposed tool was brief, easy to complete and suitable for use in a variety of clinical settings and languages.

We first conducted a review of the literature to identify comparable patient reported outcomes in communication research and clinical practice. This included examination of PROs designed for use in aphasia (language impairment), motor speech (including dysarthria and apraxia) and voice, and general communication impairment. This identified several tools with overlapping features and themes relevant to our own tool. However, no existing tools met all requirements for use. We then asked five persons with progressive dysarthria what aspects of communication they considered important to interrogate in a self-report tool. Combining information from existing tools and people living with progressive dysarthria, we focused on developing questions across three established domains of communication: physical functioning, emotional functioning, and functional impact of dysarthria.

The first series of 29 items was created across these three domains. Content validity was determined by asking domain specific experts (two speech pathologists, a neurologist, and a general physician) to discuss the suitability of the items. The list was then reduced to 25 items when considering essential conceptual and neurological elements. A trial version was condensed to 22 items following a final review by content experts and trial with the clinical cohorts (see Supplementary Materials S4 for trial items). Subsequent shorter versions of the tool were also tested, DIS 17-item [DIS-17], DIS 6-item [DIS-6])

### Participants

244 participants were recruited to the study (52.9% female). Cohorts included individuals with Huntington’s disease (n=45), Friedreich ataxia (n=38), Parkinson’s disease (n=31), Spinocerebellar ataxia (n=21), or head and neck cancer (n=20). Other diagnosis included Cerebellar ataxia, neuropathy, vestibular areflexia syndrome (CANVAS) or Multiple system Atrophy – Cerebellar (MSA-C) (n=19). Seventy age and sex matched controls also participated. See Table 1 for demographic and clinical information. Recruitment and testing were undertaken in Melbourne, Australia, and Tübingen, Germany. Clinical participants were excluded if they had comorbid neurological diseases known to impact communication (e.g., stroke, multiple sclerosis); clinical symptoms other than those caused by their primary disease; lacked competency in English or German (depending on site); a history of alcohol or drug abuse that required medical intervention; or a history of learning disability and/or intellectual impairment. Eligibility for inclusion as a healthy control required having no family history of a neurological disease and had unremarkable cognition, speech, and language function, as assessed by a speech pathologist. Cognition was assessed using the Montreal Cognitive Assessment (MoCA) ^33^. External validity was examined by administering the tools across languages and different disease groups. The study was approved by The University of Melbourne’s Human Ethics Review Committee (#30366) and the Calvary Bethlehem Hospital Ethics committee (#17122101).

**Table 1:** Clinical and demographic information for cohort.

|  | All (N = 244,♀= 129) | FRDA (N = 38,♀= 22) | SCA (N = 21) ♀= 10) | Other ataxias (N= 19,♀= 14) | HD (N = 45,♀= 24) | PD (N = 31,♀= 9) | H&N (N= 20,♀= 5) | HC (N=70,♀= 45) | Kruskal Wallis (p) |
| --- | --- | --- | --- | --- | --- | --- | --- | --- | --- |
| Age at assessment yrs ± SD (range) | 54.4 ± 15.6 (20-88) | 44.8 ± 11.4 (20-79) | 58.5 ± 9.9 (41-72) | 59.4 ± 12.0 (31-79) | 46.4 ± 14.6 (22-68) | 68.6 ± 6.2 (57-79) | 65.8 ± 7.6 (53-88) | 52.6 ± 2.8 (20-88) | <0.001 |
| Age at onset yrs ± SD (range) | 53.1 ± 14.2 (18-87) | 43.9 ± 15.4 (18-64) | 33.2 ± 15.9 (18-59) | 48.7 ± 16.2 (25-65) | 49.6 ± 13.8 (25-67) | 54.7 ± 14.2 (18-87) | 64.0 ± 8.1 (52-87) | NA | <0.001 |
| Disease duration ± SD (range) | 8.9 ± 8.0 (0-33) | 10.3 ± 4.7 (3-15) | 16.4 ± 9.8 (18-59) | 9.1 ± 9.7 (1-29) | 5.0 ± 3.6 (1-15) | 13.9 ± 7.3 (2-33) | 1.8 ± 1.7 (0-5) | NA | <0.001 |
| Speech severity score (intelligibility) | 1.2 ± 1.2 (0-4) | 2.3 ± 0.9 (1-4) | 1.6 ± 1.1 (0-4) | 1.9 ± 0.9 (0-4) | 0.9 ± 1.0 (0-4) | 1.7 ± 1.2 (0-4) | 1.8 ± 1.3 (0-4) | 0 ± 0 (0-0) | <0.001<sup>b</sup> |
| Speech severity score (naturalness) | 1.3 ± 1.2 (0-4) | 2.3 ± 1.1 (0-4) | 1.9 ± 1.1 (0-4) | 2.0 ± 1.1 (0-3) | 1.3 ± 0.9 (0-3) | 1.9 ± 1.0 (0-4) | 1.3 ± 1.0 (0-4) | 0 ± 0 (0-0) | <0.001<sup>b</sup> |
| MoCA total score ± SD (range) | 25.8 ± 3.6 (11-30) | 25.6 ± 1.6 (23-27) | 24.2 ± 2.4 (21-27) | 23.7 ± 3.0 (19-29) | 25.4 ± 5.2 (11-30) | 25.5 ± 2.8 (19-30) | 23.8 ± 2.0 (21-28) | 27.4 ± 2.8 (22<sup>c</sup>-30) | <0.001<sup>a</sup> |

Note: HD = Huntington's disease; FRDA = Friedreich ataxia; SCA = Spinocerebellar ataxia; PD = Parkinson's disease; H&N = Head and neck patients; Other = other neurodegenerative diseases; HC = Healthy controls; yrs = years; Speech severity scored on scale of 0-4 where 0=unremarkable and 4=severe; <sup>a</sup> H&N vs HC ( $p<.001$ ), Other diagnosis vs HC ( $p=0.013$ ); <sup>b</sup> most groups differed from each other; <sup>c</sup> one 83 year old participant received a score of 14 on the MoCA and their data was removed.

### Reference tests and clinically meaningful endpoints

All participants were assessed using the Dysarthria Impact Scale (DIS) (index test). Convergent validity was examined by asking participants to also complete the Voice Handicap Index (VHI) ^34^. The VHI was used as a reference test for communication performance. The full version of the VHI is a PRO with 30 items exploring the psychosocial consequences of voice disorders. The Short Form 36 (SF-36) ^35^ was used as a reference test for general health and wellbeing. The SF-36 is a PRO examining generic health and quality-of-life. Participants also provided speech samples that were recorded for rating and analysis. Samples were acquired to investigate the link between speech production self-reported speech related quality of life. Speech was recorded using the Redenlab® Desktop or Online software in a quiet room. Speech tasks included two iterations of a sustained vowel /a:/ and an unprepared monologue for approximately one minute. Speech samples were examined perceptually by expert speech clinicians on an ordinal scale of 0-4 where 0=unremarkable and 4=severe, as per previous work ^36, 37^. Concurrent validity was examined by producing objective composite measures of intelligibility and naturalness using Redenlab’s® Analyze pipeline derived from the two tasks as per earlier work ^38, 39^ and comparing them to DIS scores.

### Demonstrating methodological quality of DIS

Methods outlined in the QUADAS-2 (Quality Assessment of Diagnostic Accuracy Studies-2) ^40^ helped optimize the clinometric properties of the DIS. These criteria were considered to minimize bias and improve applicability. The two reference tests, VHI and SF-36, and the index test (DIS), were interpreted separately (i.e., expert clinicians rating speech were blinded to DIS outcomes). The index test was administered to all participants irrespective of clinical presentation. The reference tests were applied to all participants who completed the index test where possible. The duration between the index and reference tests was brief (i.e., <24 hours) to minimize opportunities for participant performance or perception to change between tests (e.g., medication for PD can influence performance over the day). We recruited four different clinical populations to ensure a diverse representative sample of dysarthria was exposed to the index test. Data are presented in detail to adequately describe specific aspects of the index test. The index test can easily be reproduced based on information provided in this manuscript. Lastly, reasonable definitions of normal/abnormal performance on the index and reference tests are provided.

### Statistical analysis

Statistical analyses were conducted using SPSS (IBM SPSS Statistics for Windows, Version 28.0. Armonk, NY: IBM Corp). Construct validity was determined by comparing data from three distinct reference tools (the VHI (voice QoL) ^34^, and the SF-36 (general health QoL) ^35^. Receiver operating characteristic (ROC) curves and associated statistics were used to calculate the sensitivity and specificity of DIS, against the reference tests, together with 95% confidence intervals. Cut-off thresholds were chosen as 1 standard deviation below the mean for healthy controls, which gave greater weight to sensitivity, without minimizing the importance of specificity or losing much area under the curve (AUC). Sensitivity scores provided an estimate of the proportion of participants who were identified as presenting with reduced quality of life due to impaired speech. Specificity calculations estimated the proportion of unimpaired participants who were identified as not presenting with reduced QoL. Differences between disease groups were explored with plots and summary statistics. Pearson’s correlations between DIS and clinically relevant metrics such as disease and dysarthria severity and dysarthria were calculated to explore these relationships.

### Test-retest reliability

Intra individual stability and reliability for the DIS was explored by examining agreement between scores provided one month apart. This short duration was considered long enough to wash out some familiarity effects and brief enough to ensure the disease had not progressed. Agreement was examined using a Bland–Altman plot in addition to correlation, as it was anticipated both samples would be strongly correlated ^41^. Only data from the first assessor were included in the final validation data analysis. Repeated assessments were only used for establishing reliability, as determined in our pre-analysis plan.

### Item reduction

The original list of items in DIS included 22 questions. An iterative approach was adopted to reduce the number of items with the aim of retaining accuracy but reducing completion time and burden on participants. This could then yield a complete set of questions, in addition to a brief version, for use in time poor clinical settings. At each step, the correlation between the total DIS and each item was calculated, and the item with the weakest correlation (or more than one item if similarly weak) was eliminated (see Supplementary Materials S1). Sensitivity, specificity, and AUC were calculated at each step to help determine the main possibilities for a shorter questionnaire. Principal component analysis (PCA) was initially performed but it resulted in the same item reduction.

### Calculating Minimal Detectable Change (MDC) and Minimal Clinically Important Difference (MCID)

MDC was estimated via Within-Subject Standard Deviation (WSSD) values. WSSD was calculated to quantify individual-level test–retest variability for the DIS-17 and DIS-6. The standard deviation of the differences between test and retest scores (SD_diff) was first derived from repeated assessments conducted one month apart in the ataxia subgroup. The WSSD was then computed using the standard formula:

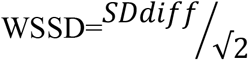

In the context of test–retest or within-subject variability calculations, the square root of 2 is used to adjust for the variance of the difference, which is twice the within-subject variance.

MCID reflects a patient-perceived threshold of benefit and is ideally estimated using an external anchor to captures what patients judge to be a meaningful change. In this study we used MCID was estimated using a distribution-based approach, because no anchor (e.g. patient-reported global change) was available. Here we used the widely accepted “0.5 × SD” approach ^42^, which estimates MCID as half of the standard deviation of the group score distribution.

### Translation

Materials were translated from English into German, French, Polish, Czech, Portuguese and Turkish (see Supplementary Materials S2 for alternative language versions). Translation protocols are described in Kraus et al ^43^ and included both forward and backward translations required for each language.

## Results

Participant groups varied in terms of speech impairment severity, age at assessment, age at onset, disease duration and MoCA scores (see Table 1).

### Relationship between the Dysarthria Impact Scale and reference tests

The index test (DIS) scores were compared to reference tests (SF-36 and VHI) as well as associated clinical features to determine validity of the tool. All versions of the DIS (DIS 22-item [DIS-22], DIS 17-item [DIS-17], DIS 6-item [DIS-6]) were strongly correlated with the reference tests as well as expert blinded consensus ratings of dysarthria (see Tables 2 and 3, Figure 1 and Supplementary Materials S5 for details). DIS scores did not appear to be associated with disease duration or cognition.

**Table 2:**
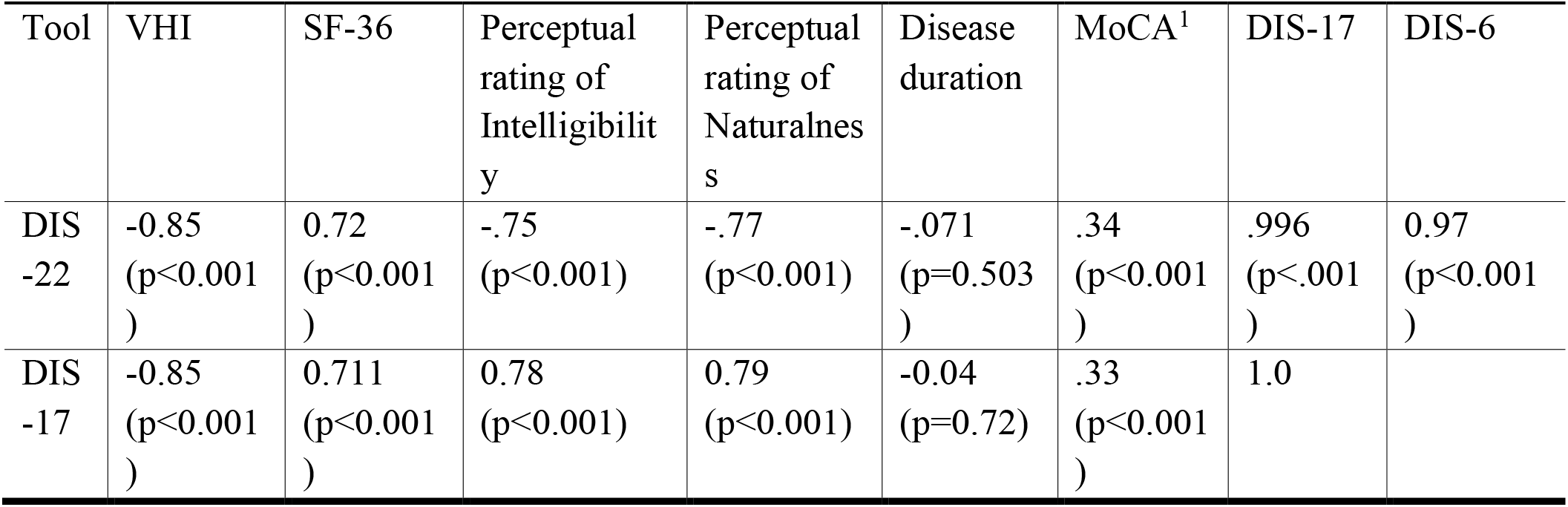
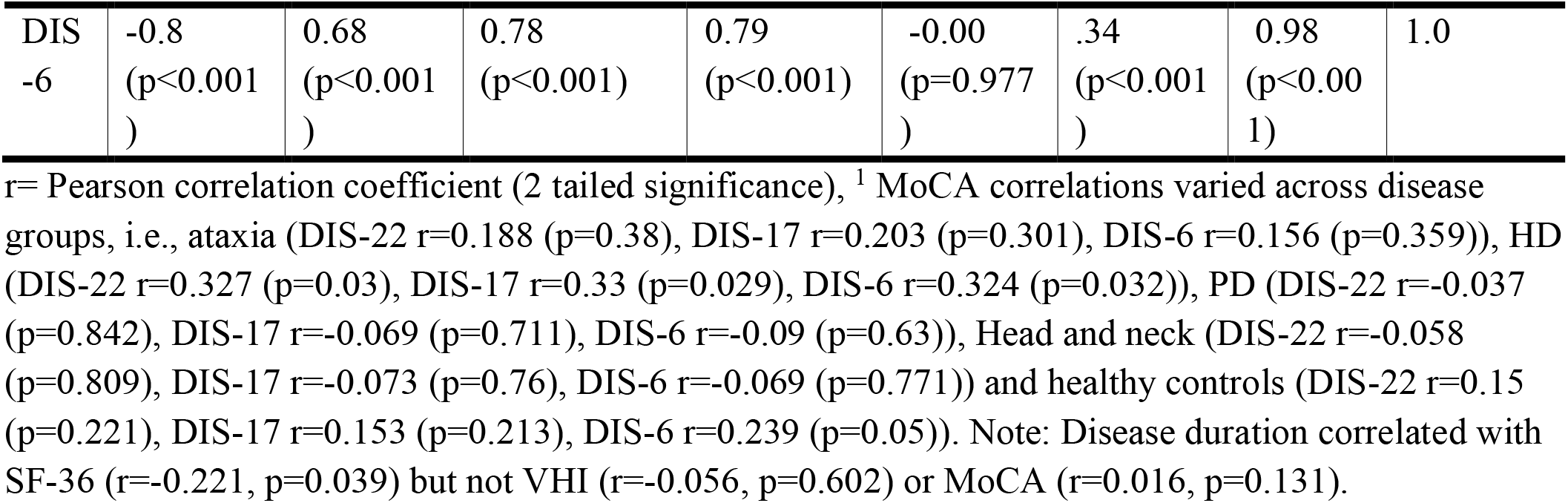
Relationship between DIS-22, DIS-17 and DIS-6 and reference tests.

**Table 3:** Relationship between perceptual dysarthria ratings and DIS scores.

| Perceptual feature and scale (consensus) | DIS-17 | DIS-6 |
| --- | --- | --- |
| Intelligibility rating (Likert) | -0.75 (p<0.001) | -0.75 (p<0.001) |
| Naturalness rating (Likert) | -0.77 (p<0.001) | -0.76 (p<0.001) |
| Intelligibility rating (DME) | 0.78 (p<0.001) | 0.76 (p<0.001) |
| Naturalness rating (DME) | 0.79 (p<0.001) | 0.77 (p<0.001) |

Note: Likert scale ranges from 0-4 with 0 representing unremarkable and 4 representing severe. DME= direct magnitude estimation with 100 representing mild dysarthria and lower values representing more severe dysarthria

**Figure 1:**
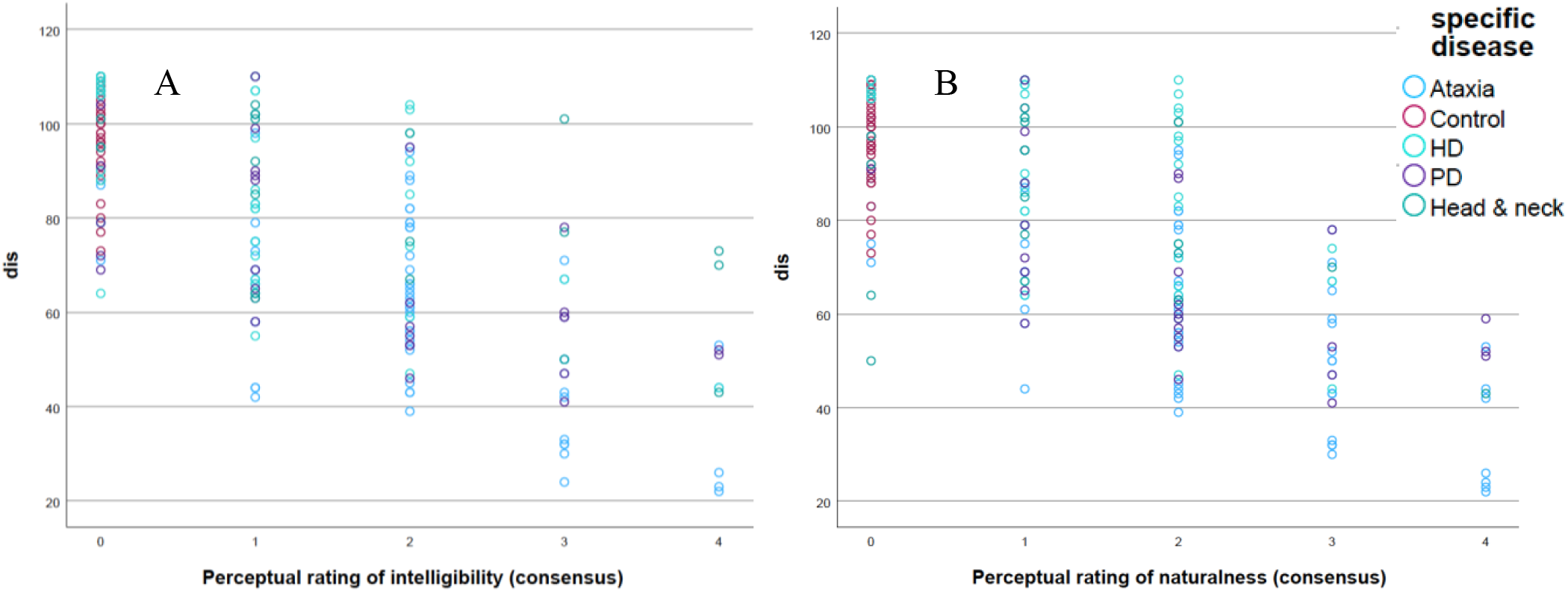
Association between DIS-22 and perceptual (A - intelligibility) and (B – naturalness) by disease group. Note HD= Huntington’s disease, PD=Parkinson’s disease, Ataxia refers to all dominant and recessive ataxias.

### Item reduction

The twenty-two item DIS was tested and validated in the entire cohort (all participants). Two shorter versions of the DIS were produced following item reduction. Following removal of redundant items based on statistical item by item comparisons, the Full Version of DIS contained 17 items, from the original 22 items. A six item Brief Version of the DIS (DIS-6) was also developed using the iterative correlation approach described in the ‘item reduction’ section in the methods. See Supplementary Materials (S1) for a breakdown of the iterative item reduction rounds and corresponding AUCs. Both revised versions of the DIS and their translations into Spanish, French, German, Polish, Czech and Turkish are available in the Supplementary Materials S2.

### Sensitivity and specificity analysis

The sensitivity and specificity of three versions of the DIS (DIS-22, DIS-17, DIS-6) is described in Table 4 and Figure 2. DIS-22 and DIS-17 yielded very similar AUC with equivalent sensitivity and specificity as described in the Statistical analysis section of the methods (i.e., cut-off thresholds were chosen as 1 standard deviation below the mean for healthy controls). The shortest version of DIS (DIS-6) yields 5% less sensitivity than the longer versions (0.93 – 0.88 = 0.05), potentially not correctly identifying 5 people in 100 with speech related quality of life issues. It, however, has 2% higher specificity than longer versions of the questionnaire.

**Table 4:** Area under the curve, sensitivity, and specificity of DIS versions.

| Test version | Area of Curve | SE | *Asymptotic Significance* | *Asymptotic 95% CI (lower – upper boundaries)* | *Sensitivity* | *Specificity* |
| --- | --- | --- | --- | --- | --- | --- |
| DIS-22 | 0.971 | 0.012 | <0.001 | 0.948-0.994 | 0.93 | 0.84 |
| DIS-17 | 0.963 | 0.015 | <0.001 | 0.933-0.993 | 0.93 | 0.84 |
| DIS-6 | 0.96 | 0.016 | <0.001 | 0.923-0.986 | 0.88 | 0.86 |

**Figure 2:**
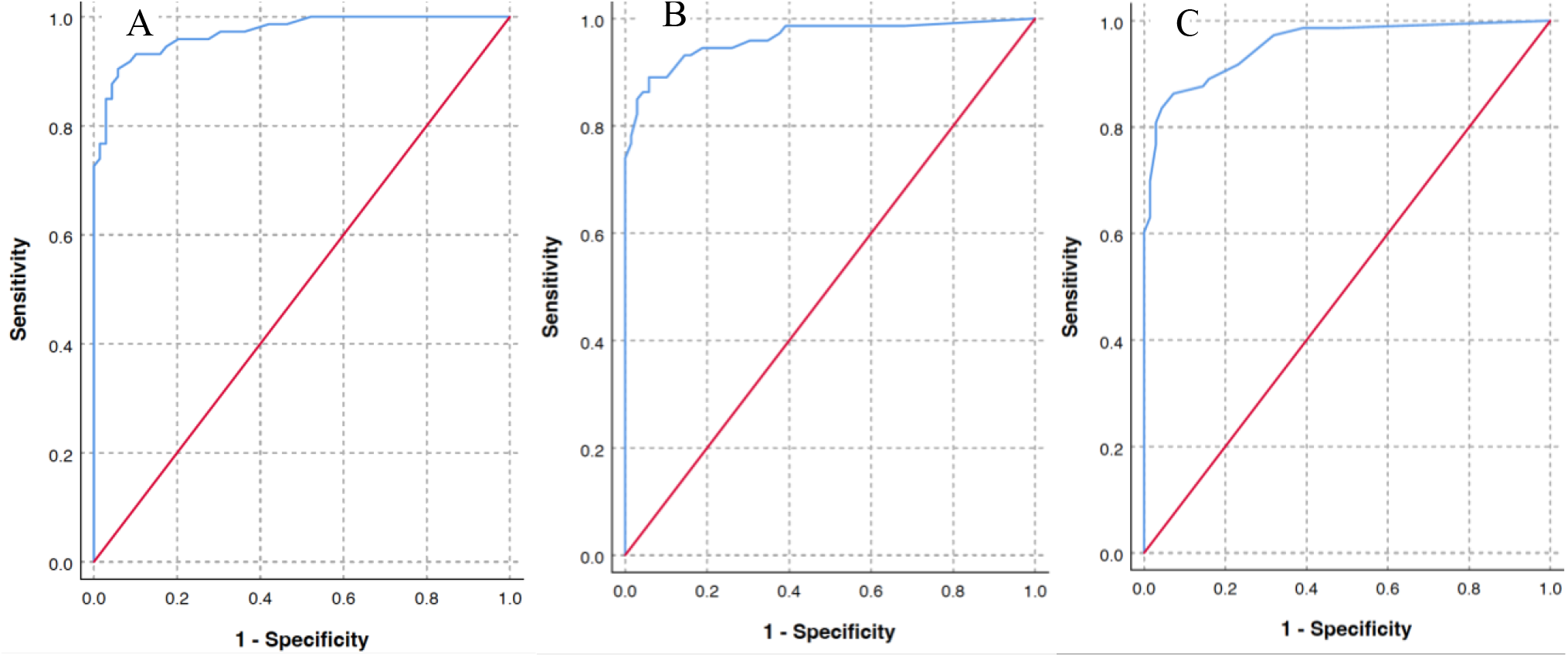
ROC curves for DIS versions. Note: A= DIS-22; B= DIS-17; C= DIS-6

### Performance of clinical and control groups on DIS-6 and DIS-17

A test of between-subject effects using Univariate Analysis of Variance revealed a difference across diseases for the DIS-17 (F=47.71, p<.001) and the DIS-6 (F=42.57, p<.001) (see Table 5), indicating that both versions of the DIS can capture (i.e. are sufficiently sensitive to) differences in speech related quality of life resulting from diverse disease groups. Where dysarthria is commonly worse (e.g., Parkinsons and ataxia, speech related quality of life is worse, compared to healthy controls and groups where dysarthria is less severity (e.g., Huntington’s disease). A post hoc analysis was run comparing the VHI and DIS-6 and DIS-17 using ANOVA with Diagnosis as the predictive factor. VHI does not appear to distinguish between HD and PD, whereas the DIS places HD tangibly (and significantly) higher as estimated by F scores (See Supplementary Materials Table S6).

**Table 5:** DIS scores for full and brief versions across disease groups.

|  |  | DIS-6 (Min=6, max=30) | DIS-17 (Min=17, max=85) |
| --- | --- | --- | --- |
| Diagnosis | N | Mean ± SE (95% CI) | Mean ± SE (95% CI) |
| Ataxia <sup>1</sup> | 73 | 16.3 ± 0.71 (14.91-17.7) | 47.7 ± 1.76 (44.22-51.17) |
| Huntington's disease | 45 | 23.87 ± 0.85 (22.2-25.53) | 67.98 ± 2.11 (63.85-72.14) |
| Parkinson's disease | 31 | 18.1 ± 1.02 (16.09-20.11) | 53.61 ± 2.54 (48.6-58.62) |
| Head and neck ward | 20 | 20.5 ± 1.27 (18.0-23.0) | 63.4 ± 3.17 (57.16-69.64) |
| Healthy controls | 69 | 28.11 ± 0.69 (26.74-29.47) | 79.01 ± 1.73 (75.6-82.41) |

Note: <sup>1</sup> Ataxia diagnostic group includes Autosomal recessive spastic ataxia of Charlevoix-Saguenay (ARSACS), Multiple system atrophy type C (MSA-C), Paraneoplastic cerebellar ataxia, Cerebellar Ataxia with Neuropathy and Vestibular Areflexia Syndrome (CANVAS), Idiopathic ataxia; SE= standard error; CI= confidence interval; Min=minimum; Max=maximum.

### Test-retest reliability

DIS was repeated in the same group of participants (N=71, ataxia only) one month apart. Test-retest scores are presented in Figure 3. A one sample t-test yielded a mean difference between DIS-17 iterations of -0.62 (t=-0.9, p=0.38) and -0.23 (t=*-*.*69*, p=0.49) for DIS-6, both suggesting agreement between means. The mean difference and standard deviation of each version of DIS were used to construct the limits of agreement (LoA) for Bland-Altman plots [i.e., DIS-6= -0.23 (mean difference) ±1.96 (95% of differences) *2.76 (standard deviation) = -5.63/5.27 and DIS-17= -0.62±1.96*5.88=- 12.15/10.91] see Figure 3. Approximately 5% of responses are expected to fall outside the limits of agreement. Data suggest approximately 7% of responses in this sample sit outside of the LoA. Given the retest challenge was only completed by people with ataxia, where disease is known to vary as a function of fatigue and sleep, mood and physical conditioning, the level of agreement observed here is considered satisfactory. Correlation between both versions was high (R=0.98, p<0.001).

**Figure 3:**
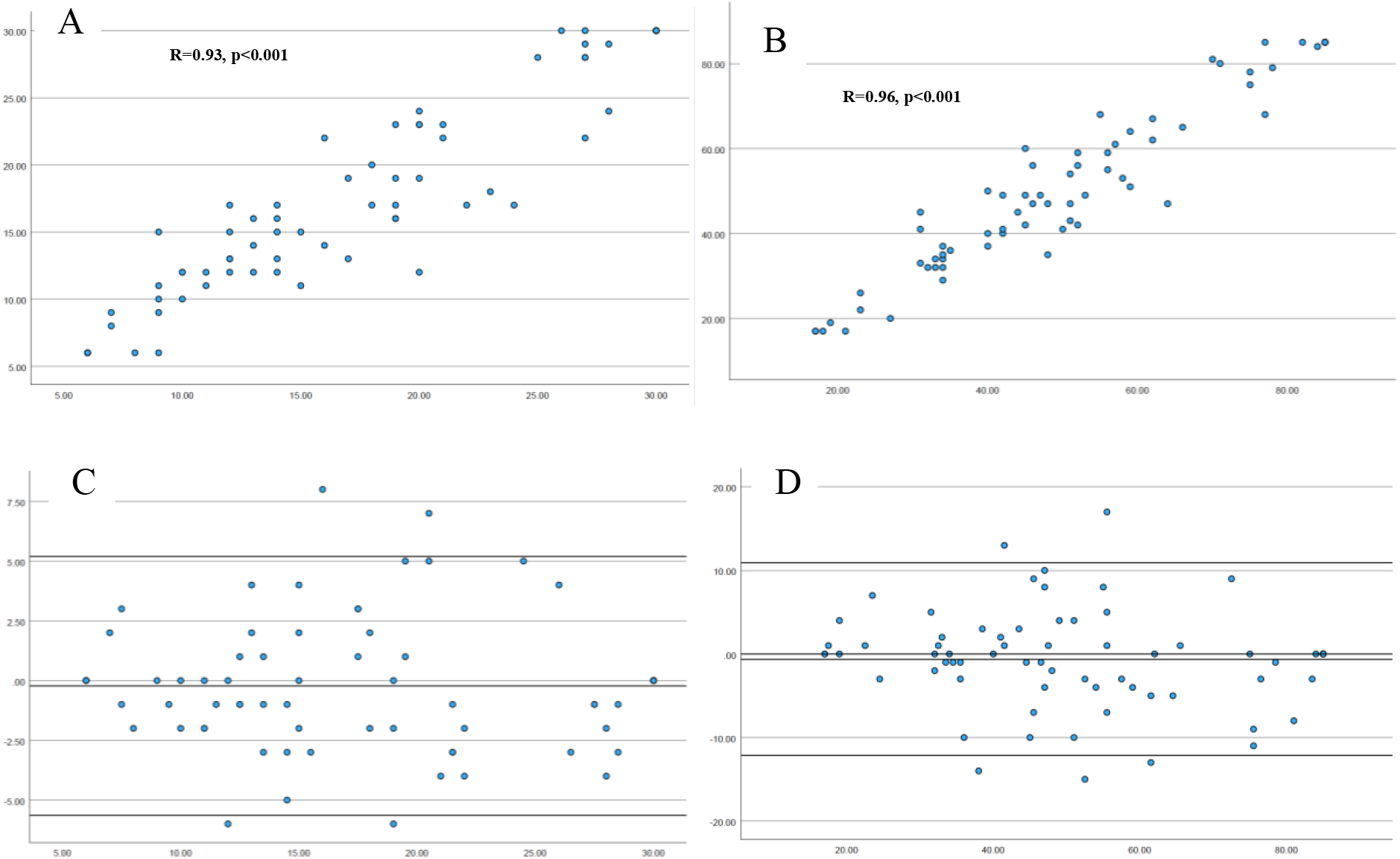
Test-retest for DIS-6 and DIS-17 (one month apart) Note: A: DIS-6 raw scores; B: DIS-17 raw scores; C: Bland-Altman of DIS-6 with LoA; D: DIS-17 with LoA.

**Figure 4.**
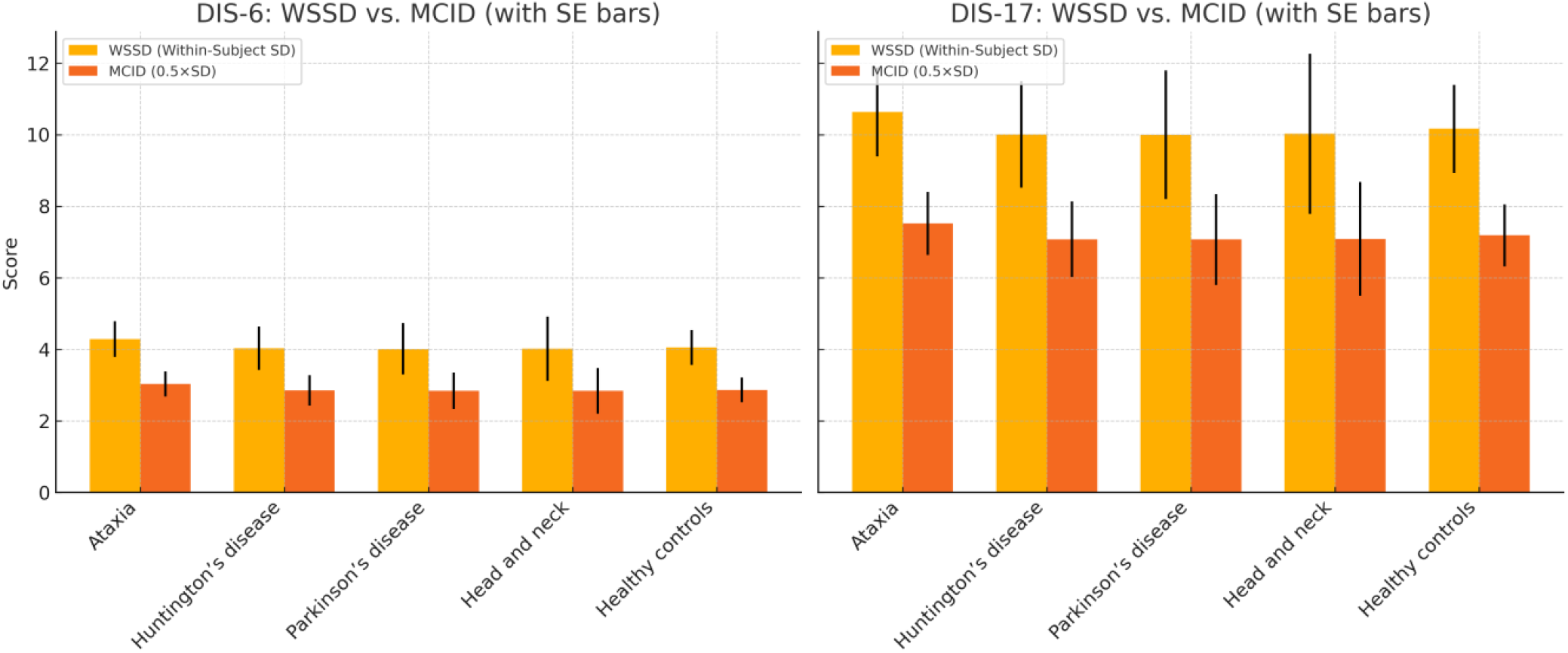
MCID and WSSD across groups. Within-subject standard deviation (WSSD) and estimated minimal clinically important difference (MCID) for DIS-6 and DIS-17 across diagnostic groups.

### MDC and MCID

Using the reported standard deviation of the difference scores, the WSSD for both the DIS-17 and DIS-6 was calculated as follows: SD_diff is the standard deviation of the differences between test and retest scores (√2 ≈ 1.414), DIS-17: SD_diff = 5.88 [WSSD = 5.88 / 1.414 ≈ 4.16] and DIS-6: SD_diff = 2.76 [WSSD = 2.76 / 1.414 ≈ 1.95]. WSSD is used here as a measure of minimal detectable change. MCIDs calculated here are distribution-based approximations and reflect the smallest change that could be considered meaningful for each group. The estimated minimal clinically important differences (MCIDs) for DIS-6 and DIS-17 ranged from approximately 5 to 15 points across diagnostic groups, providing clinically relevant thresholds to interpret meaningful change in speech-related quality of life (Figure 6 and Supplementary Materials S5).

## Discussion

The Dysarthria Impact Scale is a brief, repeatable and validated patient reported outcome for measuring the effect of neurological speech impairment on the speaker. It yields meaningful data on dysarthria in people with ataxia, Parkinson’s disease, Huntington’s disease and head and neck patients. The DIS’ utility was established using published guidance on clinical tool development, expert judgement, clinical experience, published reports and familiarity with legacy tools in the domain. It provides a sensitive and specific clinical outcome assessment that can be administered as part of experimental and clinical assessment and trial protocols, translated into 6 languages.

Historical PROs for motor speech are limited by their length (e.g., up to 160 items), complexity (e.g., positive and negative wording), limited validation data, or validation in non-dysarthria contexts (e.g., voice only). The DIS contains questions related to physical, emotional and functional performance, focused on practical scenarios where speech may impact interactions (e.g., social isolation, talking on the phone, complex conversations), indicating good construct validity.

Speech related quality life as measured by both the full and short forms of the DIS were strongly linked to overall dysarthria severity, as measured by expert speech pathologist ratings, indicating strong content and concurrent validity. More severe dysarthria is linked to lower speech related quality of life. DIS scores were also aligned with performance on the general health questionnaire (SF-36) and the voice related quality of life PRO (VHI), thus indicating good convergent construct validity. Cognition, as measured by the MoCA, had a small influence on responses to DIS, but only significantly in the HD cohort. Scores on the DIS were not directly related to disease duration in these cohorts but did vary as a function of disease itself, indicating good external validity. These data suggest that cognition and overall disease duration may contribute to QoL but are not primary drivers related to communication.

Consideration of speech as a distinct domain of assessment will enable more accurate phenotyping and acknowledgment of the relative contributions speech deficits can make to a patients’ profile.

The DIS demonstrated strong convergent validity with both the VHI (r = –0.85 for DIS-22 and DIS-17; –0.80 for DIS-6) and the SF-36 (r = 0.72 for DIS-22; 0.68 for DIS-6). These robust correlations support the DIS as a valid measure of speech-related quality of life, capturing overlapping constructs of vocal handicap and general health-related quality of life. The consistency across short and long versions reinforces its applicability in time-constrained settings. However, convergent validity may be limited by the VHI’s focus on voice rather than articulatory impairment, and the SF-36’s lack of communication-specific domains.

Perhaps surprisingly, DIS scores were not associated with disease duration (DIS-22: r = –0.071, p = .503) or global cognitive function (MoCA correlations nonsignificant in most disease groups), suggesting that the tool discriminates between speech-specific impairment and other disease features. This supports its construct specificity. However, the small but significant MoCA correlations in some subgroups (e.g., HD) suggest a potential cognitive bias in self-report that warrants further investigation in populations with more pronounced cognitive decline ^44^.

The test–retest reliability of the DIS was high across a 1-month interval (r = 0.98 for both DIS-6 and DIS-17), indicating excellent temporal stability. Bland–Altman analysis revealed acceptable limits of agreement (DIS-6: –5.63 to 5.27; DIS-17: –12.15 to 10.91), with 7% of scores falling outside these bounds. Given that this was assessed in a population (ataxia) with known day-to-day speech variability ^45^, the observed variability is considered acceptable. Future work is needed to confirm these findings across other clinical groups and longer periods.

The DIS-22 and DIS-17 exhibited comparable sensitivity (0.93) and specificity (0.84), with negligible loss of AUC (DIS-22: 0.971 vs DIS-17: 0.963). The DIS-6 retained a high AUC (0.96) with slightly reduced sensitivity (0.88) but increased specificity (0.86), suggesting it may be preferable in screening scenarios where false positives are more problematic. The minor reduction in sensitivity for DIS-6 likely reflects exclusion of items relating to complex emotional or interactional speech tasks, which may be more affected in early-stage or mild cases.

The Minimal Clinically Important Difference was not calculated using direct patient-reported anchors (such as patient global impression of change). Instead, a distribution-based method was applied to estimate a provisional MCID ^42^. According to this method, the MCID was estimated as half the standard deviation of baseline scores across clinical cohorts. This approach provides a practical and provisional clinical threshold for interpreting changes on the DIS, pending future confirmation using patient-reported anchors or longitudinal intervention studies. Comparison of within-subject standard deviation (WSSD) and minimal clinically important difference (MCID) values across diagnostic groups provides critical insight into the interpretability of score changes on the DIS. For both DIS-6 and DIS-17, WSSD values were consistently lower than corresponding MCID estimates across all disease groups, indicating that typical intra-individual variability is unlikely to obscure clinically meaningful changes. This separation between measurement noise (WSSD) and meaningful change (MCID) supports the DIS as a stable and responsive tool for monitoring treatment effects or disease progression. Interestingly, WSSD values were similar across groups, whereas MCID values were more variable, reflecting differences in overall speech-related burden and score dispersion between conditions. For example, ataxia and Parkinson’s disease groups exhibited larger MCID values, consistent with broader variability in speech impact severity. These findings suggest that while the DIS performs robustly across populations, disease-specific benchmarks may enhance interpretability in longitudinal applications.

At a group level, mean QoL scores varied significantly across groups for both DIS-6 and DIS-17 (p < .001). Ataxia and Parkinson’s disease groups had the lowest scores (DIS-17 means: 47.7 and 53.6), indicating more impaired speech-related quality of life, aligning with known speech severity in these conditions ^36-38, 46-48^. In contrast, HD participants reported comparatively higher DIS scores (mean = 68.0), despite known speech changes in HD ^8^. This may reflect reduced insight, anosognosia, or distinct communicative priorities ^49^. Head and neck cancer patients had intermediate scores as would be expected in wards were approximately one third of patients present dysarthria ^50^, with supporting the scale’s sensitivity across etiologies. Healthy controls scored near the ceiling, confirming discriminative capability.

### Limitations

The DIS has strong psychometric properties, however this study has several limitations that warrant consideration. First, the cross-sectional design precludes conclusions about the scale’s responsiveness to disease progression or intervention. While short-term test–retest reliability was established over a one-month interval in individuals with ataxia, longer-term data are needed to assess whether the DIS can reliably detect clinically meaningful change over time. The absence of longitudinal data also limits the ability to derive robust estimates of minimal clinically important differences. Additionally, the analysis of group differences may be constrained by unequal sample sizes across diagnostic categories, which may influence statistical power and generalisability of findings. Test–retest reliability was limited to a single clinical group, further constraining inferences about stability across broader populations. While cognitive screening using the MoCA suggested minimal influence of cognition on DIS scores overall, subgroup analyses indicated mild associations in some groups, such as Huntington’s disease. This may reflect reduced insight or executive dysfunction, which could affect the reliability of self-reported outcomes. While the DIS was translated into multiple languages, psychometric evaluation was restricted to the English and German versions; further validation is required to confirm cross-cultural and linguistic equivalence. Finally, the use of reference tools such as the SF-36 and VHI, though appropriate, may not fully capture the multidimensional nature of communication-related quality of life, highlighting the need for additional anchors (e.g., patient global impression of change) in future work.

## Conclusions

Motor speech disorders can lead to dramatic reductions in quality of life ^9, 51, 52^ and quality of disease management ^53^. The burden of communication decline is dealt to the affected individual, their entire family and wider network ^54-57^. Impaired speech leads to daily disadvantage, triggering social marginalization and underemployment ^3, 58^. The DIS can quantify the deleterious consequences of dysarthria including limited activities and social participation (e.g. communicating over the telephone, talking to friends), and that these are related to key environmental factors (e.g. communicating in noisy environments). The magnitude of dysarthria’s impact on the speaker and their community means it is essential that speech related quality of life is quantified in clinical and experimental settings.

The DIS is a brief, reliable, and valid patient-reported outcome measure that captures the impact of speech impairment on quality of life across diverse neurological conditions. It demonstrates strong convergent and discriminant validity, excellent test–retest reliability, and robust sensitivity and specificity across full and abbreviated versions. The tool distinguishes between disease groups and aligns closely with perceptual ratings of dysarthria severity, supporting its utility in both clinical and research contexts. While further longitudinal validation and cross-cultural testing are needed, the DIS represents a clinically meaningful and scalable instrument for assessing speech-related quality of life in individuals with motor speech disorders.

## Data Availability

All statistical data are available in Supplementary Materials. Additional output can be requested from the authors.

## Authors’ Roles

Design: APV, LG, MS

Execution: APV, CSJC, LG, MG, MS

Analysis: APV, GH, MG, MS Draft: APV

Editing of final version of the manuscript: APV, LG, CSJC, GH, MG, MS

## Financial Disclosures and conflicts of interest

Prof Vogel and Ms. Graf are employees of Redenlab Ltd, a speech neuroscience company.

A/Prof Hepworth, Dr Chan, and Dr Gauß report no conflict of interest.

Prof Synofzik has received consultancy honoraria from Ionis, UCB, Prevail, Orphazyme, Biogen, Servier, Reata, GenOrph, AviadoBio, Biohaven, Zevra, Lilly, Quince, and Solaxa, all unrelated to the present manuscript.

**Statistical Analysis** was conducted by Graham Hepworth PhD, The University of Melbourne, Australia

## Acknowledgementss

We thank our collaborators for preparing translations of the tool including Drs Serge Pinto and Stephanie Borel (French), Drs Özgür Öztop Çakmak and Atay Vural (Turkish), Dr Sandra Rojas (Spanish), Dr Anna Sobańska (Polish), Drs Gustavo Noffs and Luciana Abranches (Portuguese), and Dr Martin Vyhnalek and Lucie Stovickova and (Czech).

## Funding Acknowledgements

This study was supported by the National Ataxia Foundation (NAF), the German Hereditary Ataxia Society (DHAG), the “Stiftung Hoffnung” (to M.S.), and the Center for Rare Diseases (ZSE), Tübingen, Germany. A.P.V. received salaried support from the National Health and Medical Research Council, Australia (#1082910) and the Australian Research Council Future Fellowship (#220100253) and received funding from the Alexander von Humboldt Foundation. L.S. and M.S. are members of the European Reference Network for Rare Neurological Diseases - Project ID No 739510. This work was supported by the European Union, project European Rare Disease Research Alliance (ERDERA), GA n°101156595, funded under call HORIZON-HLTH-2023-DISEASE-07 (to M.S.)

