## Supplementary materials for "Development and validation of the Dysarthria Impact Scale"

**S1 Steps involved in item removal**

The statistical approach for item removal is described in the Methods section of the main document. Item removal involved discarding more than one item, as before. Below are the steps taken to reduce the DIS from 22 items to 17 and then to 6. Sensitivity and specificity were chosen using mean and standard deviation SD for the control group as a cutoff.

<u>All 22 items</u>: AUC = 0.971 Sens = 0.932 Spec = 0.841
Weakest correlation with item 3 (r = 0.773), item 5 (r = 0.678), item 6 (r = 0.759), item 21 (r = 0.777), item 22 (r = 0.768).

<u>17 items</u>: AUC = 0.963 Sens = 0.932 Spec = 0.841
Weakest correlation with item 8 (r = 0.800), item 17 (r = 0.797).

<u>15 items</u>: AUC = 0.967 Sens = 0.932 Spec = 0.826
Weakest correlation with item 1 (r = 0.817), item 12 (r = 0.817), item 13 (r = 0.818).

<u>12 items</u>: AUC = 0.967 Sens = 0.918 Spec = 0.870
Weakest correlation with item 9 (r = 0.820).

<u>11 items</u>: AUC = 0.966 Sens = 0.904 Spec = 0.841
Weakest correlation with item 7 (r = 0.842), item 18 (r = 0.845), item 19 (r = 0.849).

<u>8 items</u>: AUC = 0.964 Sens = 0.877 Spec = 0.841
Weakest correlation with item 11 (r = 0.881), item 14 (r = 0.878).

6 items:	AUC = 0.955	Sens = 0.877	Spec = 0.855

Weakest correlation with item 15 ($r = 0.888$).

### S2 Dysarthria Impact Scale versions

**Dysarthria Impact Scale (FULL VERSION)** ***English***

| How would you rate the overall quality of your speech? | Very good (1) | Good (2) | Fair (3) | Poor (4) | Very poor (5) |
|---|---|---|---|---|---|

Please rate how much you agree with the following statements about your speech from "fully agree" to "do not agree at all."

| | Fully agree (1) | Agree (2) | Neither agree nor disagree (3) | Do not agree (4) | Do not agree at all (5) |
|---|---|---|---|---|---|
| 1. Because of my speech, I find it difficult to be understood in noisy environments. | | | | | |
| 2. My speech affects my social life. | | | | | |
| 3. Because of my speech, I cannot communicate to a full extent. | | | | | |
| 4. Because of my speech, I avoid talking to strangers. | | | | | |
| 5. I feel self-conscious when speaking. | | | | | |
| 6. I feel like people treat me differently because of my speech. | | | | | |
| 7. People unfamiliar with me have difficulty understanding my speech. | | | | | |
| 8. It takes a great deal of effort to make myself understood. | | | | | |
| 9. I depend on others because of my speech. | | | | | |
| 10. I feel like people consider me less competent because of my speech. | | | | | |
| 11. I am frustrated with my speech. | | | | | |
| 12. Because of my speech, I avoid talking on the phone. | | | | | |
| 13. Because of my speech, it is difficult for me to describe events. | | | | | |
| 14. I feel less competent because of my speech. | | | | | |
| 15. I am often asked to repeat myself. | | | | | |
| 16. My speech affects my ability to perform in the workplace. * | | | | | |
| 17. Because of my speech, I have difficulty participating in fast or complex conversations. | | | | | |

**In non-working participants: does their speech affect communication in a bureaucratic/business environment? e.g. banking, official necessities.* *© 2016-2025 Vogel, Graf, Synofzik*

**Dysarthria Impact Scale (DIS-6)** ***English***

| How would you rate the overall quality of your speech? | Very good (1) | Good (2) | Fair (3) | Poor (4) | Very poor (5) |
|---|---|---|---|---|---|

Please rate how much you agree with the following statements about your speech from "fully agree" to "do not agree at all."

| | Fully agree (1) | Agree (2) | Neither agree nor disagree (3) | Do not agree (4) | Do not agree at all (5) |
|---|---|---|---|---|---|
| 1. My speech affects my social life. | | | | | |
| 2. I feel isolated or lonely because of my speech. | | | | | |
| 3. People unfamiliar with me have difficulty understanding my speech. | | | | | |
| 4. Because of my speech, I avoid talking on the phone. | | | | | |
| 5. Because of my speech, it is difficult for me to describe events. | | | | | |
| 6. Because of my speech, I have difficulty participating in fast or complex conversations. | | | | | |

*© 2016-2025 Vogel, Graf, Synofzik*

Translations available in full published text or from author.

medRxiv would not allow us to publish them in languages other than English.

**Supplementary Materials Section 3**

**S3 Domain selection**

Literature and clinical experience informed selection domains relevant to the impact of dysarthria on quality life. Specific themes included functional (as used here Voice Handicap Index (VHI) [6]), social-emotional and physical functioning (as used here Voice Related Quality of Life (VRQOL)[7], and acceptance (as used here the Dysarthria Impact Profile (DIP)[8]).

**S3 Chosen domains and items**

Questions were designed to capture information across the select domains. Table S3.1 shows the first round of domains and items selected for interrogation. Successive iterations are described in Tables S3.2-3.4, showing the working progress across domains and comments made by the team. Item reduction and refinement occurred following discussion among the assembled research team.

**Table S3.1** *Iteration 1*

| functional | emotional | physical |
|---|---|---|
| 1. noisy space<br>2. providing information<br>3. describing events<br>4. turn taking<br>5. multi-speaker setting<br>6. unfamiliar listeners<br>7. asked for repetition<br>8. reduction in content<br>9. avoid talking on the phone<br>10. not participating in complex or quick conversations<br>11. find alternatives for communication, e.g., text, IM, email | 1. social setting<br>2. workplace<br>3. interacting with friends, family self-conscious<br>4. avoidance talking<br>5. dependence on others<br>6. frustration<br>7. isolation / loneliness<br>8. treat me differently<br>9. respond to me differently<br>10. self-esteem / confidence<br>11. less competent<br>12. change in identity<br>13. stigma<br>14. condescension from listener<br>15. embarrassment | 1. dysarthria severity ➔ better: dysarthria intelligibility?<br>2. speech is different at different times in the day e.g., when tired<br>3. increased effort to be understood |

**Table S3.2** *Iteration 2*

| functional | emotional | physical |
|---|---|---|
| 1. noisy space<br>2. providing information ➔ replace by #3<br>3. describing events<br>4. turn taking<br>5. multiple speaker setting ➔ same like# 4 and # 10<br>6. unfamiliar listeners | 12. social setting<br>13. workplace<br>14. interacting with friends, family self-conscious<br>15. avoidance talking ➔ is already part of #8, #9, #14<br>16. dependence on others | 27. dysarthria severity ➔ better: dysarthria intelligibility?<br>28. speech is different at different times in the day e.g., when tired<br>29. increased effort to be understood |

| | |
|---|---|
| 7. asked for repetition<br>8. reduction in content<br>9. avoid talking on the phone<br>10. not participating in complex or quick conversations<br>11. find alternatives for communication, e.g., text, IM, email | 17. frustration<br>18. isolation / loneliness<br>19. treat me differently ➔ is the same like #20, merge with #20<br>20. respond to me differently<br>21. self-esteem / confidence<br>22. less competent<br>23. change in identity ➔ similar to #21<br>24. stigma<br>25. condescension from listener ➔ is the same like #20, merge with #20<br>26. embarrassment ➔ is similar to #21, merge with #21 |

**Table S3.3** *Iteration 3*

| functional | emotional | physical |
|---|---|---|
| 1. noisy space<br>2. providing information ➔ replace by #3 AGREED<br>3. describing events<br>4. turn taking<br>5. multiple speaker setting ➔ same like# 4 and # 10 DISAGREE ON BOTH<br>6. unfamiliar listeners<br>7. asked for repetition<br>8. reduction in content<br>9. avoid talking on the phone<br>10. not participating in complex or quick conversations<br>11. find alternatives for communication, e.g., text, IM, email | 12. social setting<br>13. workplace<br>14. interacting with friends, family self-conscious<br>15. avoidance talking ➔ is already part of #8 NOPE, #9 YES IN PART – SPECIFIC SETTING, #14 SAME AS PREVIOUS POINT<br>16. dependence on others<br>17. frustration<br>18. isolation / loneliness<br>19. treat me differently ➔ is the same as #20, merge with #20 AGREED<br>20. respond to me differently<br>21. self-esteem / confidence<br>22. less competent<br>23. change in identity ➔ similar to #21 PARTLY DISAGREE<br>24. stigma<br>25. condescension from listener ➔ is the same like #20, merge with #20 – SORT OF- WE DO | 27. dysarthria severity ➔ better: dysarthria intelligibility?<br>28. speech is different at different times in the day e.g., when tired<br>29. increased effort to be understood |

| | |
|---|---|
| | HAVE SOME DUPLICATION HERE<br>26. embarrassment ➔ is similar to #21, merge with #21<br>CONCEPTUALISED DIFFERENTLY |

#### S4 Scatterplots of associations between DIS-22 and DIS-17 tests

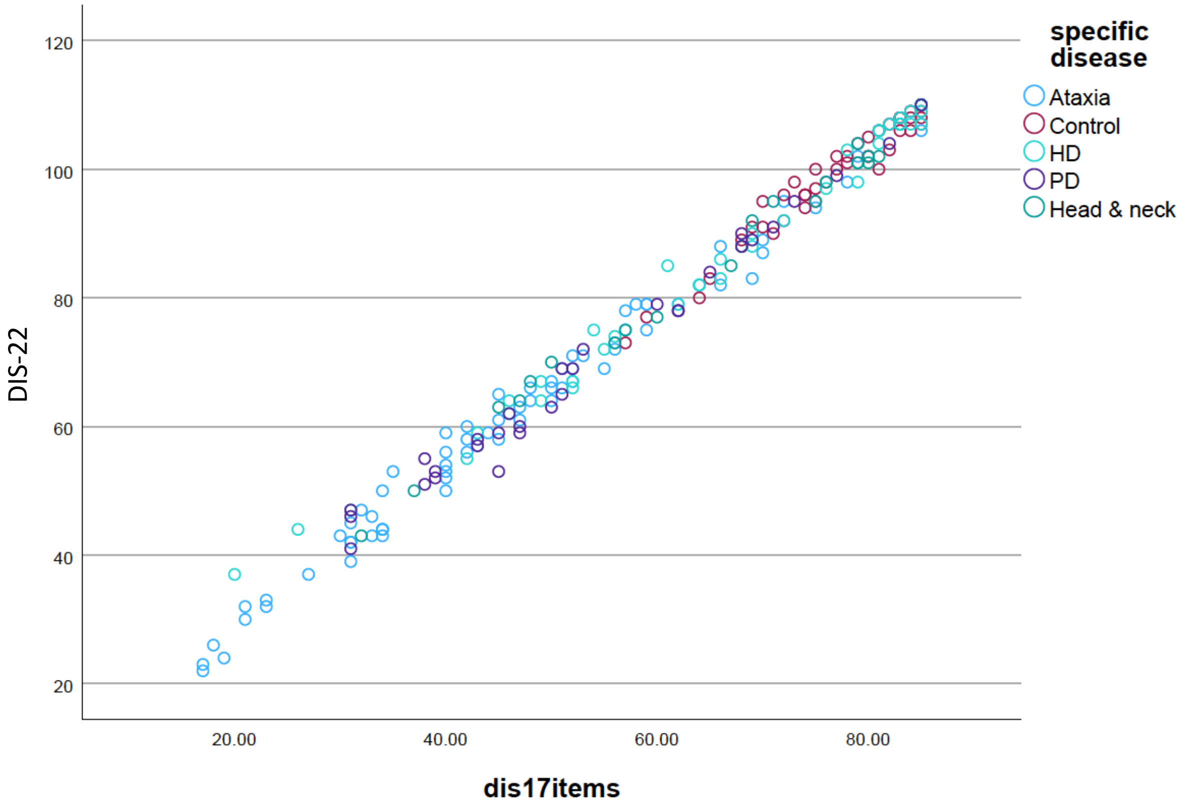

**Figure S4.1 Association between DIS-22 and DIS-17.** Note HD= Huntington's disease, PD=Parkinson's disease, and ataxia (referring to all dominant and recessive ataxias).

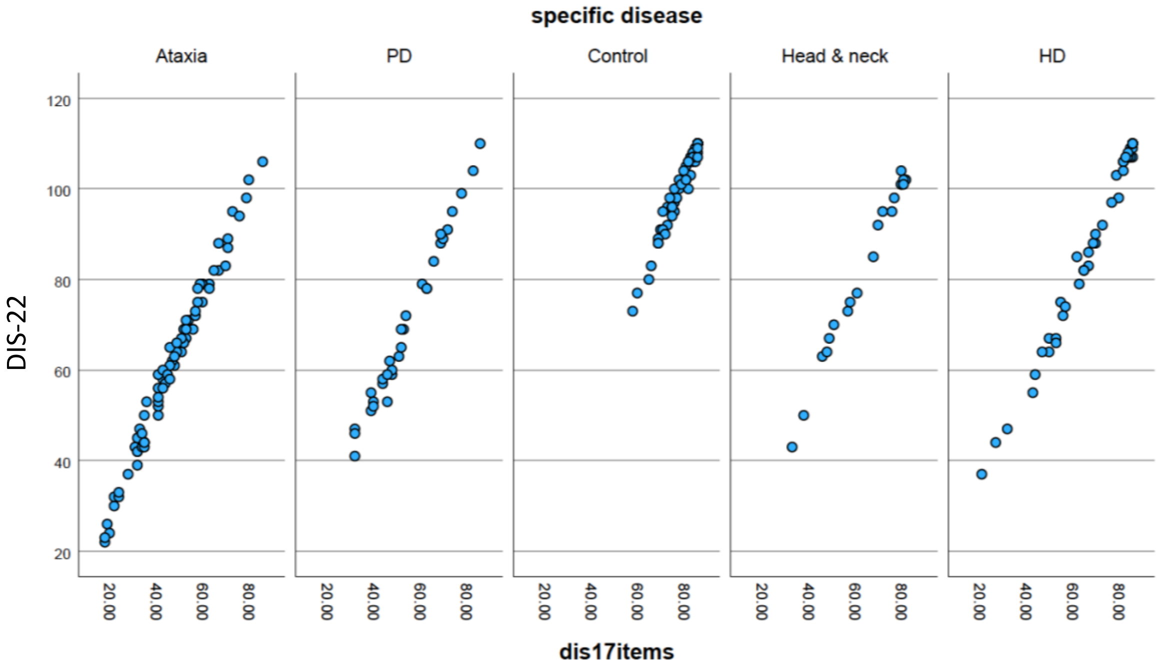

**Figure S4.2 Association between DIS-22 and DIS-17 by disease group.** Note HD= Huntington's disease, PD=Parkinson's disease, and ataxia (referring to all dominant and recessive ataxias).

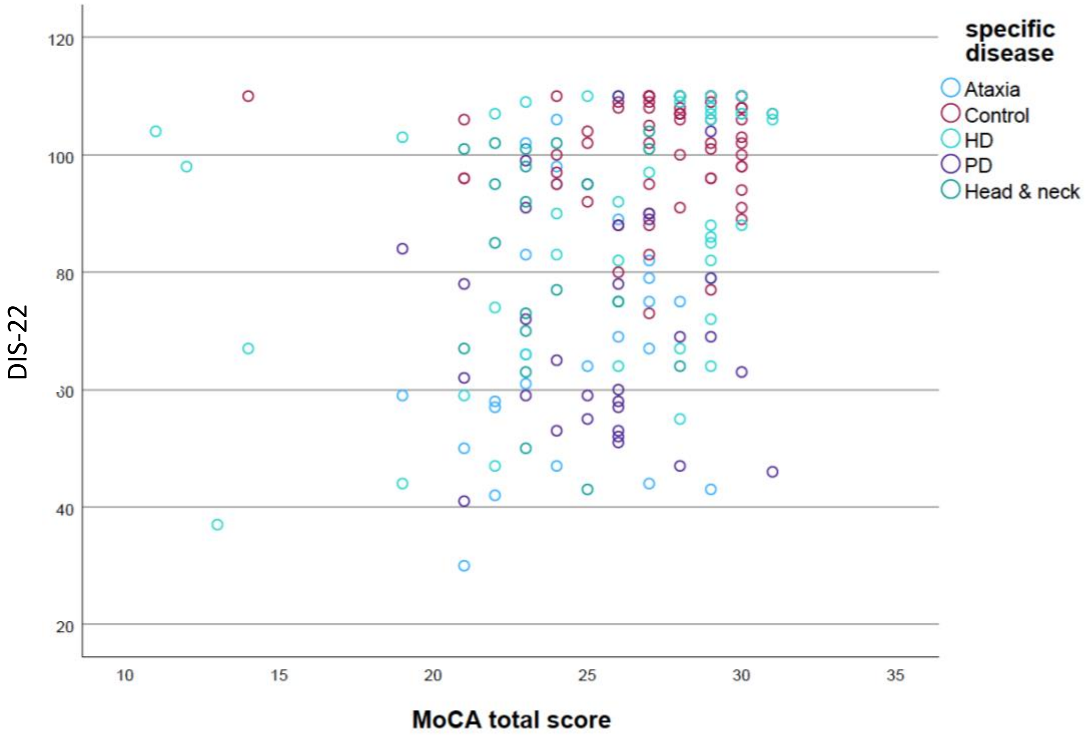

**Figure S4.3 Association between DIS-22 and MoCA by disease group.** Note HD= Huntington’s disease, PD=Parkinson’s disease, and ataxia (referring to all dominant and recessive ataxias).

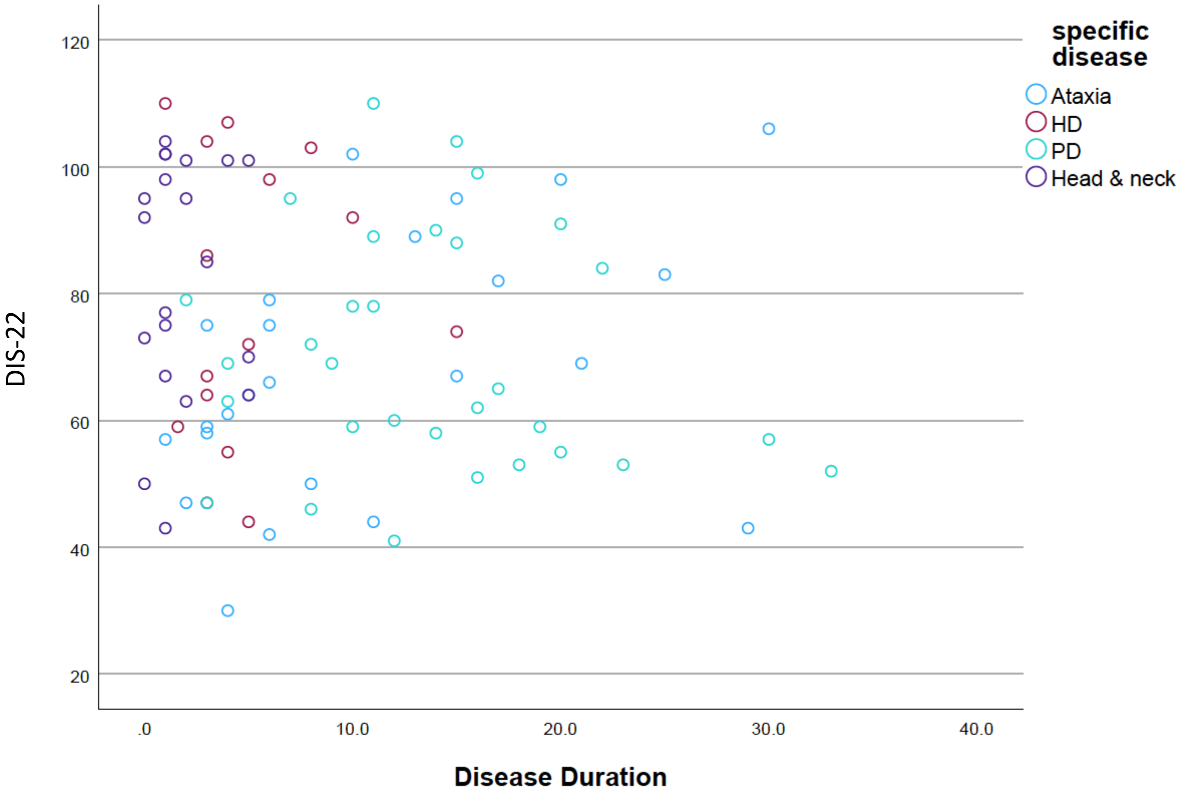

**Figure S4.4 Association between DIS-22 and disease duration by disease group.** Note HD= Huntington's disease, PD=Parkinson's disease, and ataxia (referring to all dominant and recessive ataxias).

#### S5 Minimal detectable change and minimal clinical detectable difference

##### S5.1 Table with WSSD (MDC) and MCID across groups

| Diagnosis | DIS-6 WSSD | DIS-6 WSSD SE | DIS-6 MCID (0.5√SD) | DIS-6 MCID SE | DIS-17 WSSD | DIS-17 WSSD SE | DIS-17 MCID (0.5√SD) | DIS-17 MCID SE |
|---|---|---|---|---|---|---|---|---|
| Ataxia | 4.29 | 0.5 | 6.07 | 0.36 | 10.63 | 1.24 | 15.04 | 0.88 |
| Huntington's disease | 4.03 | 0.6 | 5.7 | 0.42 | 10.01 | 1.49 | 14.15 | 1.06 |
| Parkinson's disease | 4.02 | 0.72 | 5.68 | 0.51 | 10 | 1.8 | 14.14 | 1.27 |
| Head and neck | 4.02 | 0.9 | 5.68 | 0.64 | 10.02 | 2.24 | 14.18 | 1.58 |
| Healthy controls | 4.05 | 0.49 | 5.73 | 0.34 | 10.16 | 1.22 | 14.37 | 0.86 |

**Note:** Within-subject standard deviation (WSSD) and estimated minimal clinically important difference (MCID) for DIS-6 and DIS-17 across diagnostic groups. SE = standard error, SD = standard deviation**.**

##### S5.2 Example Use Case: Evaluating Speech Intervention in Ataxia

A pharmaceutical company conducts a 12-week trial testing a novel treatment aimed at improving speech function in individuals with spinocerebellar ataxia. Participants complete the DIS-17 at baseline and again at the end of the study. The average DIS-17 score improves from 48.0 to 56.5, a mean change of +8.5 points.

Using the study's data:

- For the ataxia group, the MCID for DIS-17 is ~7.5 points, and the WSSD is ~10.6.
- This 8.5-point improvement exceeds the MCID, suggesting the change is clinically meaningful.
- The change also lies within the expected test–retest variability range (±WSSD), but with statistical confirmation (e.g., paired t-test), it provides evidence that the intervention produced a real, patient-perceived benefit.

If instead the change had been only 4 points:

- It would fall below the MCID, even if statistically significant, and may be deemed clinically negligible.
- Investigators might then conclude the treatment effect is measurable but not yet meaningful to patients.

**S6 Comparing relative difference scores between DIS-6, DIS-17 and VHI across disease groups**

| Disease Group | VHI Mean (95% CI) | DIS-17 Mean (95% CI) | DIS-6 Mean (95% CI) |
|---|---|---|---|
| Ataxia | 62.8 (57.4–68.1) | 46.1 (42.8–49.4) | 15.8 (14.5–17.1) |
| Control | 19.0 (13.7–24.4) | 79.0 (75.6–82.4) | 28.1 (26.7–29.4) |
| HD | 42.8 (36.2–49.4) | 68.0 (63.8–72.2) | 23.9 (22.2–25.5) |
| PD | 41.8 (33.7–49.9) | 53.6 (48.5–58.7) | 18.1 (16.1–20.1) |
| Head & Neck | 31.4 (21.5–41.3) | 63.4 (57.1–69.7) | 20.5 (18.0–23.0) |

Note: Group comparison (One-way ANOVA) VHI: $F(4, 228) = 33.56$, $p < .001$ | DIS-17: $F(4, 233) = 51.44$, $p < .001$ | DIS-6: $F(4, 233) = 45.95$, $p < .001$. The F tests the effect of specific disease. This test is based on the linearly independent pairwise comparisons among the estimated marginal means.

**Supplementary Materials References**

1. Ross CA, Reilmann R, Cardoso F, et al. Movement Disorder Society Task Force Viewpoint: Huntington's Disease Diagnostic Categories. Movement disorders clinical practice 2019;6:541-546.
2. Unified Huntington's Disease Rating Scale: reliability and consistency. Huntington Study Group. Movement Disorders 1996;11:136-142.
3. Penney JB, Vonsattel J-P, Macdonald ME, Gusella JF, Myers RH. CAG repeat number governs the development rate of pathology in Huntington's disease. Annals of Neurology 1997;41:689-692.
4. Paulsen JS, Langbehn DR, Stout JC, et al. Detection of Huntington's disease decades before diagnosis: the Predict-HD study. J Neurol Neurosurg Psychiatry 2008;79:874-880.
5. Group HS. Unified Huntington's Disease Rating Scale (UHDRS). Available from: www huntington-study-group org/UHDRS/tabid/67/Default aspx [Last accessed 5 June 2014] 2016.
6. Jacobson BH, Johnson A, Grywalski C, et al. The Voice Handicap Index (VHI): Development and Validation. American Journal of Speech-Language Pathology 1997;6:66-70.
7. Hogikyan ND, Sethuraman G. Validation of an instrument to measure voice-related quality of life (V-RQOL). Journal of Voice 1999;13:557-569.
8. Walshe M, Peach RK, Miller N. Dysarthria Impact Profile: development of a scale to measure psychosocial effects. International Journal of Language & Communication Disorders 2009;44:693-715.
